# Psychological resilience, coping behaviours, and social support among healthcare workers during the COVID-19 pandemic: A systematic review of quantitative studies

**DOI:** 10.1101/2020.11.05.20226415

**Authors:** Leodoro J. Labrague

**Affiliations:** College of Nursing, Sultan Qaboos University, Oman

**Keywords:** psychological resilience, coping, mental health, healthcare workers, social support, COVID-19

## Abstract

**Aim:** To appraise and synthesize studies examining resilience, coping behaviours, and social support among healthcare workers during the coronavirus pandemic.

**Background:** A wide range of evidence has shown that healthcare workers, currently on the frontlines in the fight against COVID-19, are not spared from the psychological and mental health-related consequences of the pandemic. Studies synthesizing the role of coping behaviours, resilience, and social support in safeguarding the mental health of healthcare workers during the pandemic is largely unknown.

**Evaluation:** This is a systematic review with a narrative synthesis. A total of 31 articles were included in the review.

**Key Issues:** Healthcare workers utilized both problem-centred and emotion-centred coping to manage the stress-associated with the coronavirus pandemic. Coping behaviours, resilience, and social support were associated with positive mental and psychological health outcomes.

**Conclusion:** Substantial evidence supports the effectiveness of coping behaviours, resilience, and social support to preserve psychological and mental health among healthcare workers during the COVID-19 pandemic.

**Implications for Nursing Management:** In order to safeguard the mental health of healthcare workers during the pandemic, hospital and nursing administrators should implement proactive measures to sustain resilience in HCWs, build coping skills, and implement creative ways to foster social support in healthcare workers through theory-based interventions, supportive leadership, and fostering a resilient work environment.

## Introduction

The COVID-19 pandemic is an urgent health concern worldwide that greatly affects the mental health, well-being, and possibly work effectiveness of healthcare workers. Mounting evidence indicates that healthcare workers have suffered a deterioration in their mental and psychological health during the coronavirus pandemic, with reports from individual and review studies showing higher prevalence rates of anxiety, burnout, depression, PTSD, and psychological distress among healthcare workers compared to the general public (Chew *et al*., 2020; Shechter *et al*., 2020). In a systematic meta-analysis by shain-Ripoll *et al*., (2020) the pooled prevalence rate of stress among healthcare workers during the pandemic was 40%; furthermore, 30% of healthcare workers in the pooled analysis had anxiety, 28% experienced burnout, 24% had depression, and 13% had post-traumatic stress disorder. Hence, hospital administrators should pay attention to the mental well-being of healthcare workers as poorer mental health may put them at greater risk for PTSD and even suicide (Reger *et al*., 2020).

Evidence suggests that during stressful events (including disasters, calamities, and disease outbreak), individuals are more likely to suffer adverse mental and psychological consequences when they are not equipped with sufficient levels of resilience and coping abilities (Labrague *et al*., 2018; Duncan, 2020). Support from peers, colleagues, family, and friends has also been shown to help individuals sustain emotional balance in the face of threats and stress-inducing events (Nowicki *et al*., 2020). Earlier studies conducted during other infectious disease outbreaks such as SARS, Ebola, and MERS-CoV identified a protective role for psychological resilience, coping behaviours, and social support in healthcare workers against the psychological and mental health burden of caring for infected patients (De Brier *et al*., 2020; Baduge *et al*., 2018). Studies conducted during the COVID-19 pandemic have shown a similar pattern: psychological resilience, coping behaviours, and social support safeguard mental health and well-being among healthcare workers who are on the frontlines of the fights against this deadly virus (Labrague & De los Santos, 2020; Blanco-Donoso *et al*., 2020; Chew *et al*., 2020).

Despite the abundance of empirical studies on the topic, no studies have systematically synthesized and integrated the results. A broader perspective on the topic of protective factors for psychological and mental health among healthcare workers is vital for the formulation of effective organizational strategies to better support the mental health of healthcare workers on the frontlines of the COVID-19 pandemic. Hence, this systematic review was conducted to synthesize and integrate evidence pertaining to healthcare workers’ psychological resilience, coping behaviours, and social support during the coronavirus pandemic.

## Methods

### Design

This is a systematic review with a narrative synthesis with results reported according to the Preferred Reporting Items for Systematic Reviews and Meta-Analyses (PRISMA) protocol.

### Data Sources and Search Strategies

Relevant studies were identified through electronic database searches using PubMed, CINAHL, SCOPUS, MEDLINE, and PsychINFO from August 2020 to October 2020. The following MeSH and search terms (‘psychological resilience’, ‘psychological adaptation’ OR ‘coping’, ‘mental health’, ‘health personnel’ OR ‘healthcare workers’, ‘social support’, and ‘2019-nCoV’ OR ‘COVID-19’ OR ‘SARS-CoV-2’ OR ‘severe acute respiratory syndrome coronavirus 2’) were used individually and in combination using Boolean operators (AND, OR and NOT). In addition, cited literature in the articles reviewed were also checked for potentially relevant studies **(Figure 1)**.

**Figure 1.**
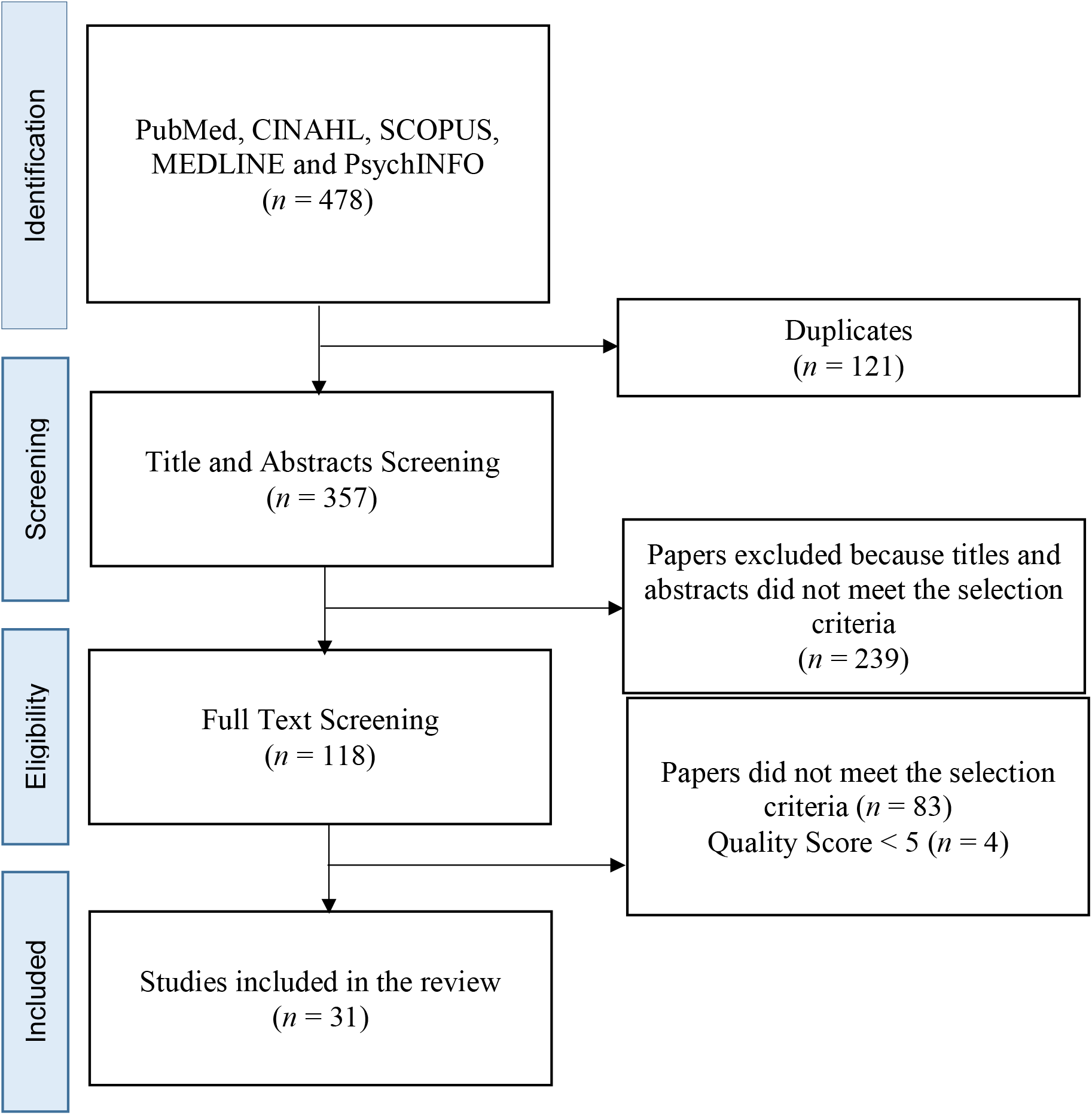
Diagram of the process used to identify references for the review.

### Inclusion Criteria

This review included primary studies assessing psychological resilience, coping, and social support among healthcare professionals during the COVID-19 pandemic. The inclusion criteria were as follows: study participants were healthcare workers, study was peer-reviewed, published since the onset of the pandemic, and published in the English language. In this review, healthcare workers are defined as people who work in healthcare settings to provide healthcare services to patients – including doctors, nurses, midwives, nursing assistants, radiologists, physiotherapists, pharmacists, healthcare assistants, and psychologists. Only studies with quantitative designs were included in this review to facilitate homogeneity of the included papers.

### Search Outcomes

The initial search yielded 478 articles, from which 121 duplicates were removed. After the removal of duplicates, 357 residual references were reviewed for relevance based on the title and abstract. After screening titles and abstracts based on the inclusion criteria, 239 articles were excluded, resulting in 118 articles. After full text reading of the articles, 87 articles were excluded due to various reasons (e.g., used different study participants, irrelevant to the objective, did not meet the eligibility criteria, and poor quality score (<5). Finally, a total of 31 articles were deemed relevant to the review. The data abstraction process is shown in **Figure 1**.

### Appraisal of Methodological Quality

Two independent researchers appraised the quality of the articles using the Joanna Briggs Institute (JBI) Critical Appraisal for Analytical Cross-Sectional Studies in order to avoid selection bias. The JBI appraisal checklist consisted of eight items examining inclusion criteria, subjects and settings, measurement exposure, use of objective and standard criteria for measurement conditions, confounding variables, management of confounding variables, outcomes measurement, and data analysis. Studies that fulfilled at least five assessment criteria were included in the review.

### Data Extraction and Synthesis

Extraction and appraisal of data was completed by two independent researchers. Using a data matrix template, the following data were extracted from the studies: authors, year of publication, country, research approach, samples, measures, key findings, and quality score (Table 1). Due to heterogeneity in the scales used and in the findings of the included studies, statistical pooling was not possible. As such, we used a narrative synthesis to describe the findings. In particular, constant comparison analysis (Miles & Huberman, 1994) was performed to compare findings across studies and to identify patterns and commonalities between studies.

**Table 1.** Summary of Included Studies.

| Authors | Country | Research Design | Samples | Measures | Key Findings | Quality Score |
| --- | --- | --- | --- | --- | --- | --- |
| Blanco-Donoso *et al.*, (2020) | Spain | Cross-sectional | 228 HWCs | SSW | 13.9% of the variance in secondary traumatic stress measure was explained by lack of staff and supervisor support.<br>Lower levels of support from co-workers amplify the negative effect of social pressure from work on traumatic stress.<br>Social pressure from work, high doses of exposure to suffering, lack of personnel and personal protective equipment, and minimal supervisor support were significant in explaining traumatic stress. | 7/8 |
| Bozdag and Ergun (2020) | Turkey | Cross-sectional | 214 HCWs | BRS; MSPSS | Mean scale score in the BRS was 18.43 out of 30.<br>Higher levels of quality of sleep, positive affective state, age and life satisfaction raised the level of psychological resilience.<br>Higher negative affective state and being a doctor meant lower psychological resilience level. | 7/8 |
| Babore *et al.*, (2020) | Italy | Cross-sectional | 595 HCWs | COPE | Lower positive attitude, higher social support, working with COVID-19 patients and higher avoidance strategies predicted higher levels of distress. | 6/8 |
| Cai (2020) | China | Cross-sectional | 534 medical | RD-CBQ | Coping strategies utilized by HCWs | 7/8 |
|  |  |  | staff |  | used strict protective measures, knowledge of virus prevention and transmission, social isolation measures, and positive self-attitude.<br>• The following provided psychological benefit in HCWs: the availability of strict infection control guidelines, specialized equipment, recognition of their efforts by hospital management and the government. |  |
| Chew *et al.*, (2020) | China | Cross-sectional | 274 resident physicians | COPE | • Stress was positively predicted by the use of avoidance as a coping strategy.<br>• Stress was negatively predicted by the use of positive thinking.<br>• Traumatic stress was positively predicted by use of avoidance as a coping strategy.<br>• The use of problem solving and use of social support as coping strategies were negative and positive predictors of traumatic stress. | 7/8 |
| Chen *et al.*, (2020) | China | Cross-sectional | 92 nurses | SIQ | • HCWs utilized the following adaptation approaches: communication with family, learning about the disease, communication with colleagues and teamwork.<br>• Least influential coping were as follows: lack of support and understanding from family and relatives; lack of protective supplies; lack of social support and recognition for medical workers and unfamiliar with special work environments,<br>working routine and use of equipment. | 78 |
| Dong *et al.*,<br>(2020) | China | Cross-sectional | 4618 (doctors,<br>nurses,<br>technician,<br>health<br>administrators<br>) | RD-CBQ | Medical staff without emotional problems were significantly more likely to cope by “adhering to infection control procedures,” “just accepting the risks,” “keeping a positive mind-set,” “keeping a healthy lifestyle,” “avoiding thinking about the risks,” “avoiding traveling,” and less “taking vitamins, herbs, or other complementary substances”.<br>Family relationships had a direct negative effect on emotional-distress levels. | 7/8 |
| Di Monte *et al.*, (2020) | Italy | Cross-sectional | 102 general practitioners | CISS; RS | Emotional exhaustion was positively correlated with emotion-oriented coping and negatively with task-oriented coping.<br>Depersonalization correlated positively with emotion-oriented coping and avoidance-oriented coping and negatively with task-oriented coping.<br>Personal Accomplishment scale was correlated negatively with emotion-oriented coping and positively with task-oriented coping.<br>Resilience had a significant positive correlation with the personal accomplishment subscale and a negative correlation with emotional exhaustion and Depersonalization subscales. | 7/8 |
| Giusti *et al.*, (2020) | Italy | Cross-sectional | 330 (doctors, nurse, nurse assistant physiotherapy) | RD-CBQ | Age, occupation, being home, work hours, psychological comorbidities, contact with COVID-19 patients, fear of infection, support from family and support from friends predicted burnout due to COVID-19. | 6/8 |
| Hou *et al.*, (2020) | China | Cross-sectional | 528 HCWs | CSCQ | PTSD symptoms were positively associated with negative coping and fatigue.<br>Negative coping moderated the relationship between self-efficacy and PTSD symptoms.<br>Negative coping also moderated the direct effect of self-efficacy on fatigue. | 6/8 |
| Huffman *et al.*, (2020) | USA | Cross sectional | 720 HCWs | CD-RISC | Resilient HCWs reported less fatigue, insomnia, stress, and anxiety than non-resilient HCWs. | 8/8 |
| Huang *et al.*, (2020a) | China | Cross sectional | 377 HCWs | CD-RISC | Psychological resilience was protective for the development of anxiety<br>83.8% of HCWs had higher psychological resilience<br>16.2% of HCWs had low psychological resilience | 6/8 |
| Huang *et al.*, (2020) | China | Cross sectional | 600 medical staff | CD-RISC | Mean scale score of the CD-RISC was 65.76 out of 100.<br>Stress score, female, less knowledge of COVID-19, less knowledge of COVID-19 protective measures, and lack of protective materials in the hospital were important related factors<br>for resilience of the medical staff. | 7/8 |
| Khalaf *et al.*,<br>(2020) | Egypt | Cross sectional | 170 physicians | BRCS | • The BRCS score was 13.45.<br>• 50% of physicians were low resilient copers, 30% were medium resilient copers and approximately 20% were high resilient copers.<br>• Gender, marital status, academic degree, specialty, years of experience, living with vulnerable family members and chronic diseases did not predict BRCS score.<br>• Psychological resilience had significant and negative correlation with depression, anxiety and stress. | 7/8 |
| Labrague & De los Santos<br>(2020a) | Philippines | Cross sectional | 325 nurses | BRCS;<br>PSSQ | • Resilience, social support, and organisational support in front-line nurses were moderate.<br>• Social support, personal resilience, and organisational support predicted COVID-19 anxiety. | 6/8 |
| Labrague & De los Santos<br>(2020b) | Philippines | Cross-sectional | 736 nurses | BRCS;<br>PSSQ | • Hospital nurses had higher scores on social support, personal resilience, and perceived general health measures than public health nurses.<br>• Personal resilience predicted dysfunctional anxiety related to coronavirus. | 6/8 |
| Lin *et al.*,<br>(2020) | China | Cross-sectional | 114 (nurses, doctors, medical staff) | CD-RISC;<br>SCSQ | • HCWs had a high level of resilience (67.04).<br>• Active coping (26.61) score was higher than the score of passive coping (10.32).<br>• Nurses obtained a lower resilience | 7/8 |
|  |  |  |  |  | score compared to other professions.<br>Moreover, active coping, depression, anxiety and mental health training were significant predictors of resilience. |  |
| Luceno-Moreno *et al.*, (2020) | Spain | Cross-sectional | 1422 HCWs | BRS | Resilience is associated in a negative and significant way with post-traumatic stress, anxiety, depression.<br>The mean scale score of the BRS was 3.02 out of a possible score of 6. | 7/8 |
| Li *et al.*, (2020) | China | Longitudinal | 356 nurses | CD-RISC | Nurses with PTSD had a significantly lower resilience than those without PTSD.<br>An increase of CD-RISC score was associated with a decrease in PTSD.<br>An increase of CD-RISC score was associated with decreased PTSD symptoms. | 6/8 |
| Mosheva *et al.*, (2020) | Israel | Cross-sectional | 1106 (physicians) | CD-RISC | Psychological resilience was negatively associated with anxiety in HCWs. | 6/8 |
| Maraqa *et al.*, (2020) | Palestine | Cross-sectional | 430 (physicians, nurses, and other allied health professionals; lab and radiology technicians) | RD-CBQ | The following coping approaches were identified by HCWs: prayers, sports, and exercise as the most common (80.5%); having clear guidelines for infection prevention (64.7%); availability of PPE (57.3%); and the support of colleagues. | 8/8 |
| Maiorano *et al.*, (2020) | Italy | Cross-sectional | 140 (physicians, nurses) | DRS; CSES | Coping strategies, especially stop unpleasant emotions and thoughts, and hardiness are protective factors and<br>reduce the effect of stress on secondary trauma. | 8/8 |
| Mi et al., (2020) | China | Cross-sectional | 1029 HCWs | RD-CBQ | • The mean score of coping measure was 18.48 (range 6–30).<br>• The most common coping strategies were physical exercise, positive attitude, and expression feeling/emotion.<br>• Coping was negatively related to depression and anxiety. | 7/8 |
| Nie et al., (2020) | China | Cross-sectional | 263 nurses | SCSQ; PSSS | • The mean score of positive coping style and negative coping style among all frontline nurses was 1.68 and 0.97 respectively.<br>• Positive coping style and negative coping style were the risk factors of COVID-19 related stress symptoms.<br>• Seven factors associated with the presence of psychological distress: working in ED, concern for family, being treated differently, the impact of the event, negative coping style, perceived social support, precautionary measures effective. | 6/8 |
| Salman et al., (2020) | Pakistan | Cross-sectional | 398 (doctors, nurse, pharmacists) | Brief-COPE | • Most frequently adopted coping strategy was religious coping followed by acceptance and coping planning.<br>• Females were observed to have significantly higher scores for behavioural disengagement, venting and religious/spiritual coping than male respondents.<br>• Respondents belonging to 26-30 | 7/8 |
|  |  |  |  |  | years' age group reported significantly less substance use than those from 31-35 years of age.<br>• Nurses had significantly higher coping style scores on denial, substance use and behavioural disengagement than doctors. |  |
| Shechter *et al.*, (2020) | USA | Cross-sectional | 657 HCWs | RD-CBQ | • Physical activity/exercise was the most commonly endorsed behaviour (59%), followed by engaging with faith-based religion and/or spirituality (23%), yoga (25%), and/or meditation (23%), engaging with talk therapy (26%) and virtual provider support groups (16%).<br>• HCWs who screened positive for acute stress reported engaging in more coping behaviours than those who screened negative. | 8/8 |
| Tam *et al.*, (2020) | China | Cross-sectional | 1280 HCWs | CD-RISD | • Psychological distress and COVID-19 stressors were negatively correlated with resilience.<br>• Resilience partially mediated the association between institutional support and psychological distress. | 6/8 |
| Vagni *et al.*, (2020) | Italy | Cross-sectional | 121 (doctors, nurses, psychologists, healthcare assistants) | CSES-SF | • HCWs utilized focused problem solving and support as coping strategies.<br>• Blocking unpleasant emotions and thoughts strategy had a significant impact on the stress levels and the components of secondary trauma, unlike the problem-focused and social<br>support strategies. | 6/8 |
| Xiao *et al.*,<br>(2020) | China | Cross-sectional | 180 medical<br>staff | SSRS | • Social support correlated significantly with anxiety and sleep.<br>• Social support negatively affected anxiety and stress levels, and positively affected their self-efficacy. | 7/8 |
| Yoruk and<br>Guler (2020) | Turkey | Cross-sectional | 377 midwives<br>and nurses | RSA | • High psychological resilience was found to be protective against depression risk. | 6/8 |
| Zhu *et al.*,<br>(2020) | China | Cross-sectional | 79 doctors and<br>86 nurses | SSRS | • The total score of positive coping was negatively correlated with the total score of anxiety and depression. | 8/8 |

**SSW:** Social Support at Work; **BRS:** Brief Resilience Scale; **MSPSS:** Multidimensional Scale of Perceived Social Support; **COPE:** Coping Orientation to Problems Experienced; **SIQ:** Stressor and Incidence Questionnaire; **CISS:** Coping Inventory for Stressful Situations; **RS:** Resilience Scale; **CSCQ:** Simplified Coping Style Questionnaire; **CD-RISC:** Connor-Davidson Resilience Scale; **BRCS:** Brief Resilient Coping Scale; **PSSQ:** Perceived Social Support Questionnaire; **PSSQ:** Perceived Social Support Scale; **CSES-SF:** Coping Self-Efficacy Scale – Short Form; **SSRS:** Social Support Rate Scale; **RD-CBQ:** Researcher-designed Coping Behaviours Questionnaire; **DRS:** Dispositional Resilience Scale; **CSES:** Coping Self-Efficacy Scale; **RSA:** Resilience Scale for Adults

### Study Characteristics

Thirty-one articles were included in the review. A majority of studies were conducted in China (14), with the remaining studies conducted in Italy (5), Philippines (2), the United States (2), Turkey (2), Spain (2), Israel (1), Palestine (1), Pakistan (1), and Egypt (1). Sample sizes ranged from 10 to 4,618 participants. Most of the healthcare workers included in the studies were nurses, doctors, nursing assistants, midwives, radiologists, physiotherapists, pharmacists, healthcare assistants, or psychologists.

Most studies had a cross-sectional research design (*n* = 30), mostly using online surveys, and one study had longitudinal research design. Most studies utilized standardized scales to measure psychological resilience, coping skills, and social support in healthcare workers. Six studies utilized research-designed questionnaires/items to identify coping skills in healthcare workers. The Cronbach’s alphas ranged from 0.81 to 0.96 in studies that reported internal consistency.

### Methodological Quality Score

Using the Joanna Briggs Institute critical appraisal checklist, the majority of the studies were rated as moderate in quality (26/31) and five were rated high. Issues related to identification of potential confounding variables and how these confounders were managed and controlled were common in the included articles.

### Major Findings

Study results were classified into the following categories: (1) levels of resilience and coping, (2) specific coping skills, (3) coping in relation to mental health, (4) resilience in relation to mental health, (5) social support in relation to mental health, and (6) strategies to enhance resilience, coping behaviours, and social support.

### Levels of Psychological Resilience and Coping

Six studies reported data on level of psychological resilience (Bozdag & Ergun, 2020; Huang *et al*., 2020a; Huang *et al*., 2020b; Khalaf *et al*., 2020; Lin *et al*., 2020; Labrague & De los Santos, 2020a) and three studies described level of coping behaviours in healthcare workers (Lin *et al*., 2020; Nie *et al*., 2020; Mi et al., 2020). High levels of psychological resilience were reported in three studies (Bozdag & Ergun, 2020; Huang *et al*., 2020a; Lin *et al*., 2020) and moderate levels of psychological resilience were reported in four studies (Luceno-Moreno *et al*., 2020; Huang *et al*., 2020b; Khalaf *et al*., 2020; Labrague & De los Santos, 2020a). In one cross-sectional study of healthcare workers assigned to radiology units in China, online survey data indicated that 83.8% of participants reported higher psychological resilience (Huang *et al*., 2020a). Similarly, a study by Lin *et al*. (2020) reported high levels of resilience in Chinese healthcare workers; however, among them, nurses were found to have lower resilience when compared to doctors and other medical staff. In a study from Turkey, healthcare workers scored 18.43 points out of a possible 30 points on the brief resilience scale (BRS), indicating a greater capability to rebound from the adversity associated with the coronavirus pandemic. On the other hand, four cross-sectional studies reported moderate levels of psychological resilience among physicians (Khalaf *et al*., 2020), medical staff (Huang *et al*., 2020b), and hospital nurses (Labrague & De los Santos, 2020a) who were on the frontlines during the pandemic. In another study, healthcare workers in Spain obtained a mean score of 3.02 on the Brief Resilience Scale (BRS), indicating a moderate capacity to bounce back to a healthy state in the face of adversity (Luceno-Moreno *et al*., 2020).

With regards to coping mechanisms, three studies measured ways of coping among healthcare workers using the Simplified Coping Style Questionnaire (Lin *et al*., 2020; Nie *et al*., 2020) and a researcher-designed coping behaviour scale (Mi *et al*., 2020). The three studies reported higher scores for positive vs. negative coping mechanisms, suggesting that when confronted with stress-inducing events such as the COVID-19 pandemic, healthcare workers are able to utilize positive coping mechanisms.

### Specific Coping Skills

Fourteen studies identified specific coping mechanisms employed by healthcare workers during the pandemic (Blanco-Donoso *et al*., 2020; Cai, 2020; Chen *et al*., 2020; Chew *et al*., 2020; Dong *et al*., 2020; Giusti *et al*., 2020; Labrague & De los Santos, 2020; Maraqa *et al*., 2020; Mi *et al*., 2020; Nie *et* al., 2020; Salman et al., 2020; Shechter *et al*., 2020; Vagni *et al*., 2020; Xiao *et al*., 2020). Among these fourteen studies reporting specific coping styles among healthcare workers during the pandemic, eleven quantitative studies (Cai, 2020; Blanco-Donoso *et al*., 2020; Chew *et al*., 2020; Dong *et al*., 2020; Giusti *et al*., 2020; Labrague & De los Santos, 2020; Nie *et a*l., 2020; Xiao *et al*., 2020; Chen *et al*., 2020; Maraqa *et al*., 2020; Vagni *et al*., 2020) indicated that healthcare workers use support from and communication with family, friends, and colleagues as their primary coping mechanisms to manage the adverse mental health consequences of the COVID-19 pandemic. Religious coping mechanisms such as praying were reported as an important coping mechanism in three cross-sectional studies. For instance, in two separate studies involving healthcare workers in Pakistan (Salman *et al*., 2020) and Palestine (Maraqa *et al*., 2020), praying and other religious activities were the highest-ranked coping mechanisms. In the United States, where prevalence of COVID-19 is highest, frontline emergency healthcare workers identified religious coping mechanisms such as praying as one of the most important ways to combat the mental and psychological burden of the pandemic (Shechter *et al*., 2020).

Involvement in distraction activities (such as engaging in sports, exercise, music, yoga, or meditation) were also identified as an important coping mechanisms utilized by healthcare workers during the height of the pandemic (Shechter *et al*., 2020; Maraqa *et al*., 2020; Mi *et al*., 2020; Chen et al., 2020; Dong *et al*., 2020). Other coping mechanisms identified by healthcare workers included learning about COVID-19 and its prevention (Chen *et al*., 2020; Cai, 2020) and adherence to infection control guidelines (Dong *et al*., 2020; Cai 2020; Maraqa *et a*l., 2020).

### Coping in Relation to Mental Health

Nine studies described the interaction between coping skills and mental health in healthcare workers during the COVID-19 pandemic (Baboe *et al*., 2020; Chew *et al*., 2020; Di Monte *et al*., 2020; Hou *et al*., 2020; Maiorano *et al*., 2020; Mi *et al*., 2020; Nie *et al*., 2020; Vagni *et al*., 2020; Zhu *et al*., 2020;). The use of positive coping mechanisms such as seeking social support, positive thinking, and problem solving were associated with lower levels of traumatic stress, stigma (Chew *et al*., 2020), psychological distress (Baboe *et al*., 2020), stress symptoms (Nie *et al*., 2020), anxiety, and depression (Zhu *et al*., 2020; Mi *et al*., 2020). On the other hand, utilization of negative coping skills, such as avoidance, were strongly linked with increased levels of emotional stress (Chew *et al*., 2020), PTSD symptoms (Hou *et al*., 2020), psychological distress (Nei *et al*., 2020; Baboe *et al*., 2020), and fatigue (Hou *et al*., 2020). In one study, the use of emotion-centred and avoidant coping styles were associated with increased levels of emotional exhaustion, while problem-centred coping styles were strongly associated with decreased scores on the depersonalization subscale and increased scores on the personal accomplishment subscale of the MBI (Di Monte *et al*., 2020). Interestingly, unlike previous studies, a two separate studies in Italy found that the use of a negative coping style – specifically, the blocking of unpleasant emotions and thoughts – was found to effectively reduce psychological distress (Vagni *et al*., 2020) and PTSD (Maiorano *et al*., 2020). Vagni *et al*. (2020) and Maiorano *et al*., (2020) both argued that by blocking negative emotions, healthcare workers are able to continue their work and experience lower perceived levels of stress.

### Resilience in Relation to Mental Health

A number of papers examined the effects of psychological resilience on the mental health of healthcare workers (12/31) (Di Monte *et al*., 2020; Huang *et al*., 2020a; Khalaf *et al*., 2020; Labrague & De los Santos, 2020a; Labrague & De los Santos, 2020b; Li *et al*., 2020; Lin *et al*., 2020; Lucero-Moreno *et al*., 2020; Maiorano *et al*., 2020; Mohave *et al*., 2020; Tam *et al*., 2020; Yoruk & Guler, 2020). Of these, eight studies reported a protective role of psychological resilience against coronavirus-related anxiety. Increased psychological resilience in healthcare workers was associated with lower incidence of pandemic-related anxiety among nurses working in hospitals (Labrague & De los Santos, 2020a) and public health centres (Labrague & De los Santos, 2020b). Results obtained from Israel and Egypt showed a similar pattern in which lower levels of coronavirus-related anxiety were associated with higher levels of resilience (Mosheva *et al*., 2020; Khalaf *et al*., 2020). In two separate studies from China, healthcare workers with higher scores on psychological resilience measures reported significantly lower levels of anxiety than those who obtained lower scores on psychological resilience measures (Huang *et al*., 2020a; Lin *et al*., 2020). In a study involving 720 healthcare workers in the United States, resilient participants were more likely than non-resilient participants to report reduced levels of anxiety, stress, fatigue, and insomnia (Huffman *et al*., 2020).

Four studies reported a strong link between personal resilience and depression (Luceno-Moreno *et al*., 2020; Lin *et al*., 2020; Khalaf *et al*., 2020; Yoruk & Guler, 2020), suggesting that interventions to enhance resilience among healthcare workers may help prevent or reduce the occurrence of depression in this population during the COVID-19 pandemic. In addition to depression and anxiety, a few more studies confirmed the protective role of psychological resilience against psychological stress (Tam *et al*., 2020; Luceno-Moreno *et al*., 2020; Khalaf *et al*., 2020), emotional exhaustion (Di Monte *et al*., 2020), and PTSD symptoms (Maiorano *et al*., 2020; Lucero-Moreno *et al*., 2020; Li *et al*., 2020). In one study, resilience partially mediated the association between institutional support and coronavirus-related distress (Tam *et al*., 2020).

### Social Support in Relation to Mental Health

Seven studies explored the causal relationship between social support and mental health outcomes in healthcare workers during the pandemic (Blanco-Donoso *et al*., 2020; Chew *et al*., 2020; Dong *et al*., 2020; Giusti *et al*., 2020; Labrague & De los Santos, 2020; Nie *et al*., 2020; Xiao *et al*., 2020). Mental health outcomes examined in relation to social support included traumatic stress, emotional distress, psychological distress, burnout, anxiety, and stress. Adequate managerial and supervisorial support and support extended by colleagues, peers, friends, and family were associated with reduced levels of traumatic stress (Blanco-Donoso *et al*., 2020; Chew *et al*., 2020) and emotional distress (Dong *et al*., 2020).

In an online cross-sectional study involving nurses in China, higher perceptions of social support explained significant variance in the psychological distress measure (Nie *et al*., 2020), while in Italy, healthcare workers who perceived greater support from family and friends reported a significant reduction in burnout symptoms (Giusti *et al*., 2020). A study involving Filipino nurses showed a similar pattern: Frontline nurses who perceived higher social support were less likely to demonstrate dysfunctional anxiety related to the coronavirus (Labrague & De los Santos, 2020). In addition, adequate social support for healthcare workers was associated with a significant reduction in stress and an improvement in self-efficacy during the pandemic (Xiao *et al*., 2020).

### Strategies to enhance resilience, coping behaviours, and social support

Several recommendations to enhance resilience, coping behaviours, and social support in HCWs were offered in the literature. Many authors suggested the development and implementation of interventions geared towards enhancing resilience in HCWs through evidence-based education and training to strengthen HCWs’ defenses against various mental and psychological consequences of the pandemic (Blanco-Donoso *et al*., 2020; Babore *et al*., 2020; Dong *et al*., 2020; Shanafelt *et al*., 2020; Labrague & De los Santos, 2020b). A few authors suggested individual and group skill training programs to foster resilience and coping skills in HCWs including online cognitive behaviour therapy or mindfulness-based therapy (Dong *et al*., 2020; Shanafelt *et al*., 2020; Giuti *et al*., 2020). Huffman *et al*., (2020) suggested the implementation of mindfulness-based stress reduction and cognitive framing to improve coping abilities in HCWs, while Maiorano *et al*., (2020) identified hardiness training to effectively enhance the ability of HCWs to withstand the burden of the pandemic and cope effectively with the stress associated with it. In addition, reinforcement of positive coping strategies through coping skills trainings were seen beneficial for strengthening the psychological well-being of healthcare providers during the pandemic (Mi *et al*., 2020; Di Monte *et al*., 2020; Khalaf *et al*., 2020).

Effective leadership was seen as vital in the promotion of mental health in HCWs and in the promotion of a resilient work environment (Chen *et al*., 2020; Shechter *et al*., 2020). By being attentive to the psychological, mental, and psychosocial needs of the HCWs hospital administrators can effectively offer support and foster resilience and coping (Blanco-Donoso *et al*., 2020; Chew *et al*., 2020; Dong *et al*., 2020). Effective leadership and organisational support through the implementation of a safe and resilient work environment, provision of complete and quality PPE and supplies to prevent infection, provision of updated and evidence-based guidelines for infection prevention, provision of accurate and timely information regarding the disease and implementation of trainings relevant to COVID-19 were seen vital to support the needs of HCWs and improve their mental well-being (Labrague & De los Santos, 2020a; Maraqa *et al*., 2020).

## Discussion

This systematic review is the first to examine psychological resilience, social support, and coping behaviours among healthcare workers during the COVID-19 pandemic and their effects on mental and psychological health. Despite the threat caused by the new virus and the pandemic’s mental health consequences, HCWs reported having moderate to high levels of psychological resilience. In explaining this occurrence, it is important to note that the studies included in the review were conducted during the first wave of the pandemic. Therefore, HCWs are still equipped with substantial personal resources (e.g., coping, self-efficacy, resilience) to combat the psychological burden caused by the coronavirus pandemic (Chen & Bonanno, 2020). However, with the increasing cases of infected patients, increasing patients’ death, increased workload due to increasing COVID-19 admissions, and lack of personal protective equipment, HCWs’ resilience, or ability to bounce back from stressful events, may eventually decline or deteriorate in the long run (Ferreira *et al*., 2020). Additionally, the lack of social connectedness and the seemingly no definite end in sight for social restrictions may contribute to this decline. As the virus continues to spread along with the threat of the new COVID-19 variants, it is imperative that proactive measures to sustain resilience in HCWs are continuously instituted. These measures may include limiting shift hours, providing adequate hospital supplies, providing rest areas in the hospitals, and providing timely updates and accurate information to HCWs regarding the virus.

Coping strategies – that is, mechanisms that an individual can employ to manage the impacts of potential threats– has been long considered an important personal resource to effectively reduce the impact of stress and its accompanying adverse consequences (Lazarus & Folkman 1987). It was evident in this review that healthcare workers utilized both positive (e.g., use of social support and praying) and negative (e.g., use of distraction activities) coping strategies to effectively manage the stress associated with the COVID-19 pandemic. Interestingly, the use of religious coping mechanisms – such as reading the Bible for Christians or reciting the Quran for Muslims – has been identified as an effective strategy to reduce stress, anxiety, and their adverse effects during the height of the pandemic. As a coping strategy, prayer provides context, social connection, and inner strength, making an individual capable of managing stress more effectively. This type of coping is not only practical, but also safe during the pandemic as it doesn’t require contact with someone. A substantial amount of studies have established a positive link between religious coping mechanisms and reduced anxiety, aggression, psychological distress, and depressive symptoms as well as enhanced optimism, hope, quality of life, and psychological health (Solaimanizadeh *et al*., 2020; O’Brien *et al*., 2019).

Seeking social support as a means of coping with adversity has been categorized as a problem-focused coping strategy (Samios *et al*., 2020) and has been found to effectively reduce stress. Mounting evidence has strongly linked adequate support from managers, co-workers, family, and friends with positive mental health outcomes for both healthcare and non-healthcare professionals during stressful and traumatic events such as calamities, accidents, disasters, and disease outbreaks (Baduge *et al*., 2018; Labrague *et al*., 2018). During the pandemic, when stress and anxiety are elevated, adequate social support may help healthcare workers maintain healthy emotional states. However, the different restrictions to combat the virus, including social distancing, lockdown, and quarantine measures, may prevent HCWs from engaging in activities previously learned to effectively cope with stress. For instance, studies conducted before the pandemic identified social support (from friends, peers, family, and even the community) and involvement of outdoor distraction activities (e.g., outdoor exercise) as important coping skills to combat stress among nurses (Lim *et al*., 2010; Ha & Sung, 2018). However, with the on-going pandemic, utilising these coping strategies can be more challenging. Because promoting social connectedness is of vital importance (as social isolation is what makes this crisis unique compared to other crises), it is essential to find creative ways to foster relationships (e.g., online social connection), to ensure that HCWs are socially and emotionally connected with their families and friends, without the risk of being infected or of infecting them. Other alternative ways to effectively cope with the mental health burden of the pandemic included formulating a new routine that incorporates healthy and optimistic behaviours, such as exercising, journaling, and writing in a gratitude journal (Stuart *et al*., 2021; Huang *et al*., 2020).

Psychological resilience, like social support, has long been considered a protective factor against the adverse psychological effects of stressful or traumatic situations (Hart, Brannan, & De Chesnay, 2014). In the context of pandemic, a wide range of evidence has demonstrated that resilient healthcare workers are more likely to rebound effectively and endure the pandemic-associated psychological burden than non-resilient healthcare workers (Foster *et al*., 2020). The role of psychological resilience in protecting individuals against the mental health consequences of an emergency or disaster situation has also been confirmed in previous studies (Labrague *et al*., 2018; Duncan, 2020). Our finding also add support to earlier research conducted prior to the pandemic in which higher resilience in healthcare workers was strongly linked to reduced burnout, compassion fatigue, anxiety, depression, and psychological distress (Mealer, Jones, & Meek, 2017).

### Limitations of the study

Although this study provided current understanding of resilience, coping behaviours, and social support among HCWs, a few limitations of the review were identified. Potentially relevant research published in other languages were excluded as this review included only articles published in English language. Further, it is worth noting that this review included articles published during the first wave of the pandemic, therefore, on-going investigations are needed to explore how resilience, coping behaviours, and social support among HCWs change through the different waves of the pandemic.

### Implications for Nursing Management

This review suggests that building resilience and increasing coping skills and social support among healthcare workers may protect them against the adverse mental and psychological health consequences of the coronavirus pandemic. As such, hospital administrators should foster psychological resilience and reinforce positive coping strategies among healthcare workers by implementing theory-tested interventions or programs. Due to restrictions including social distancing and lockdown measures, these interventions could be delivered in innovative ways, such as webinars, online workshops, and on-demand videos. Interprofessional, web-based nightly debriefing programs (Azizoddin *et al*., 2020) and online cognitive behavioural therapy (Weiner *et al*., 2020) have been demonstrated to enhance resilience and morale in healthcare workers and improve clinical processes for quality patient care. Furthermore, increasing social support may provide a sense of greater emotional security among healthcare workers, thereby reducing their apprehensions and anxiety so they can function effectively during the pandemic. If healthcare workers are encouraged to express their feelings and concerns and openly discuss their experiences and challenges in the care and management of COVID-19 patients, their morale will improve and their mental health will be sustained.

As positive coping strategies were seen to improve mental health in healthcare workers, providing training in the development of self-efficacy and effective coping skills may help healthcare workers better manage the increased work pressures that have accompanied the COVID-19 pandemic. Hospital administrators should consider increasing healthcare workers’ access to mental health professionals during the pandemic in to support their mental health needs.

## Conclusions

The review findings suggest that healthcare workers manage their stress during the height of the COVID-19 pandemic by utilizing both problem-focused (e.g., use of social support and religious practice) and emotion-focused (e.g., use of diversionary activities) coping strategies. Furthermore, this review found substantial evidence on the value and effectiveness of coping mechanisms, psychological resilience, and social support in preserving the mental health and psychological well-being of healthcare workers during disease outbreaks such as the coronavirus pandemic. Considering the global extent of the pandemic, this review is of interest to international readers – particularly hospital administrators.

## Data Availability

Data is available upon request.

## References

Azizoddin, D. R., Vella Gray, K., Dundin, A., & Szyld, D. (2020). Bolstering clinician resilience through an interprofessional, web-based nightly debriefing program for emergency departments during the COVID-19 pandemic. Journal of Interprofessional Care, 1–5.

Babore, A., Lombardi, L., Viceconti, M. L., Pignataro, S., Marino, V., Crudele, M., … & Trumello, C. (2020). Psychological effects of the COVID-2019 pandemic: Perceived stress and coping strategies among healthcare professionals. Psychiatry Research, 293, 113366.

Baduge, M. S. P., Morphet, J., & Moss, C. (2018). Emergency nurses’ and department preparedness for an ebola outbreak: A (narrative) literature review. International Emergency Nursing, 38, 41–49.

Bozdağ, F., & Ergün, N. (2020). Psychological Resilience of Healthcare Professionals During COVID-19 Pandemic. Psychological Reports, Ahead of Print.

Cai, Y. (2020). Psychological Impact and Coping Strategies of Frontline Medical Staff in Hunan Between January and March 2020 During the Outbreak of Coronavirus Disease 2019 (COVID 19) in Hubei, China. Med Sci Monit, 26, e924171.

Chew, Q. H., Chia, F. L. A., Ng, W. K., Lee, W. C. I., Tan, P. L. L., Wong, C. S., … & Phua, E. J. (2020). Perceived Stress, Stigma, Traumatic Stress Levels and Coping Responses amongst Residents in Training across Multiple Specialties during COVID-19 Pandemic—A Longitudinal Study. International Journal of Environmental Research and Public Health, 17(18), 6572.

Chen, H., Sun, L., Du, Z., Zhao, L., & Wang, L. (2020). A cross-sectional study of mental health status and self-psychological adjustment in nurses who supported Wuhan for fighting against the COVID-19. Journal of Clinical Nursing, Ahead of Print.

De Brier, N., Stroobants, S., Vandekerckhove, P., & De Buck, E. (2020). Factors affecting mental health of health care workers during coronavirus disease outbreaks (SARS, M.R. & COVID-19): A rapid systematic review. Plos one, 15(12), e0244052.

Di Monte, C., Monaco, S., Mariani, R., & Di Trani, M. (2020). From Resilience to Burnout: psychological features of Italian General Practitioners during COVID-19 emergency. Frontiers in Psychology, 11, 2476.

Dong, Z. Q., Ma, J., Hao, Y. N., Shen, X. L., Liu, F., Gao, Y., & Zhang, L. (2020). The social psychological impact of the COVID-19 pandemic on medical staff in China: A cross-sectional study. European Psychiatry, 63(1), E65.

Duncan, D. L. (2020). What the COVID-19 pandemic tells us about the need to develop resilience in the nursing workforce. Nursing Management, 27(3), 22– 27.

Folkman, S., & Lazarus, R. S. (1980). An analysis of coping in a middle-aged community sample. Journal of Health and Social Behavior, 219–239.

Foster, K., Roche, M., Giandinoto, J., & Furness, T. (2020). Workplace stressors, psychological well-being, resilience, and caring behaviours of mental health nurses: A descriptive correlational study. International Journal of Mental Health Nursing, 29(1), 56– 68.

Giusti, E. M., Pedroli, E., D’Aniello, G. E., Badiale, C. S., Pietrabissa, G., Manna, C., … & Molinari, E. (2020). The psychological impact of the COVID-19 outbreak on health professionals: a cross-sectional study. Frontiers in Psychology, 11.

Hart, P. L., Brannan, J. D., & De Chesnay, M. (2014). Resilience in nurses: An integrative review. Journal of Nursing Management, 22(6), 720–734.

Hou, T., Dong, W., Zhang, R., Song, X., Zhang, F., Cai, W., … & Deng, G. (2020). Self-efficacy and fatigue among health care workers during COVID-19 outbreak: A moderated mediation model of posttraumatic stress disorder symptoms and negative coping. Preprint

Huang, L., Wang, Y., Liu, J., Ye, P., Chen, X., Xu, H., … & Ning, G. (2020). Factors influencing anxiety of health care workers in the radiology department with high exposure risk to COVID-19. Medical science monitor: international medical journal of experimental and clinical research, 26, e926008–1.

Huang, L., Wang, Y., Liu, J., Ye, P., Cheng, B., Xu, H., … & Ning, G. (2020). Factors Associated with Resilience Among Medical Staff in Radiology Departments During the Outbreak of 2019 Novel Coronavirus Disease (COVID-19): A Cross-Sectional Study. Medical Science Monitor, 26.

Kackin, O., Ciydem, E., Aci, O. S., & Kutlu, F. Y. (2020). Experiences and psychosocial problems of nurses caring for patients diagnosed with COVID-19 in Turkey: a qualitative study. International Journal of Social Psychiatry, Ahead of Print

Khalaf, O. O., Khalil, M. A., & Abdelmaksoud, R. (2020). Coping with Depression and Anxiety in Egyptian Physicians during COVID-19,.Pandemic Pre-print

Labrague, L. J., & De los Santos, J. A. A. (2020). COVID-19 anxiety among front-line nurses: Predictive role of organisational support, personal resilience and social support. Journal of Nursing Management, 28(7), 1653–1661.

Labrague, L. J., & De los Santos, J. A. A. (2020b). Prevalence and predictors of coronaphobia among frontline hospital and public health nurses. Public Health Nursing, Ahead of Print

Langford, C. P. H., Bowsher, J., Maloney, J. P., & Lillis, P. P. (1997). Social support: A conceptual analysis. Journal of Advanced Nursing, 25(1), 95– 100.

Li, X., Zhou, Y., & Xu, X. (2020). Factors associated with the psychological well-being among front-line nurses exposed to COVID-2019 in China: A predictive study. Journal of Nursing Management. Ahead of Print

Lin, J., Ren, Y., Gan, H., Chen, Y., Huang, Y., & You, X. (2020). Factors Influencing Resilience of Medical Workers from Other Provinces to Wuhan Fighting Against 2019 Novel Coronavirus Pneumonia. Ahead of Print

Liu, Q., Luo, D., Haase, J. E., Guo, Q., Wang, X. Q., Liu, S., … & Yang, B. X. (2020). The experiences of health-care providers during the COVID-19 crisis in China: a qualitative study. The Lancet Global Health. Ahead of Print

Maiorano, T., Vagni, M., Giostra, V., & Pajardi, D. (2020). COVID-19: Risk Factors and Protective Role of Resilience and Coping Strategies for Emergency Stress and Secondary Trauma in Medical Staff and Emergency Workers—An Online-Based Inquiry. Sustainability, 12(21), 9004.

Maraqa, B., Nazzal, Z., & Zink, T. (2020). Palestinian Health Care Workers’ Stress and Stressors During COVID-19 Pandemic: A Cross-Sectional Study. Journal of Primary Care & Community Health, 11, Ahead of Print

Mealer, M., Jones, J., & Meek, P. (2017). Factors affecting resilience and development of posttraumatic stress disorder in critical care nurses. American Journal of Critical Care, 26(3), 184–192.

Mi, T., Yang, X., Sun, S., Li, X., Tam, C. C., Zhou, Y., & Shen, Z. (2020). Mental Health Problems of HIV Healthcare Providers During the COVID-19 Pandemic: The Interactive Effects of Stressors and Coping. AIDS and Behavior, 1–10.

Miles, M. B., and M. Huberman. 1994. Qualitative Data Analysis: A Sourcebook of New Methods. 2d Edition. Beverly Hills, CA: Sage Publications.

Mosheva, M., Hertz-Palmor, N., Dorman Ilan, S., Matalon, N., Pessach, I. M., Afek, A., … & Gothelf, D. (2020). Anxiety, pandemic-related stress and resilience among physicians during the COVID-19 pandemic. Depression and anxiety, 37(10), 965–971.

Nie, A., Su, X., Zhang, S., Guan, W., & Li, J. (2020). Psychological impact of COVID-19 outbreak on frontline nurses: A cross-sectional survey study. Journal of clinical nursing.

Nowicki, G. J., Ślusarska, B., Tucholska, K., Naylor, K., Chrzan-Rodak, A., & Niedorys, B. (2020). The severity of traumatic stress associated with COVID-19 pandemic, perception of support, sense of security, and sense of meaning in life among nurses: research protocol and preliminary results from Poland. International journal of environmental research and public health, 17(18), 6491.

O’Brien, B., Shrestha, S., Stanley, M. A., Pargament, K. I., Cummings, J., Kunik, M. E., … & Amspoker, A. B. (2019). Positive and negative religious coping as predictors of distress among minority older adults. International journal of geriatric psychiatry, 34(1), 54–59.

Salman, M., Raza, M. H., Mustafa, Z. U., Khan, T. M., Asif, N., Tahir, H., … & Hussain, K. (2020). The psychological effects of COVID-19 on frontline healthcare workers and how they are coping: a web-based, cross-sectional study from Pakistan. Pre-print

Samios, C., Catania, J., Newton, K., Fulton, T., & Breadman, A. (2020). Stress, savouring, and coping: The role of savouring in psychological adjustment following a stressful life event. Stress and Health, 36(2), 119–130.

Shechter, A., Diaz, F., Moise, N., Anstey, D. E., Ye, S., Agarwal, S., … & Claassen, J. (2020). Psychological distress, coping behaviors, and preferences for support among New York healthcare workers during the COVID-19 pandemic. General hospital psychiatry, 66, 1–8.

Serrano-Ripoll, M. J., Meneses-Echavez, J. F., Ricci-Cabello, I., Fraile-Navarro, D., Fiol-deRoque, M. A., Pastor-Moreno, G., … & Gonçalves-Bradley, D. C. (2020). Impact of viral epidemic outbreaks on mental health of healthcare workers: a rapid systematic review and meta-analysis. Journal of affective disorders, 277, 347–357.

Solaimanizadeh, F., Mohammadinia, N., & Solaimanizadeh, L. (2020). The relationship between spiritual health and religious coping with death anxiety in the elderly. Journal of religion and health, 59(4), 1925–1932.

Tam, C. C., Sun, S., Yang, X., Li, X., Zhou, Y., & Shen, Z. (2020). Psychological Distress Among HIV Healthcare Providers During the COVID-19 Pandemic in China: Mediating Roles of Institutional Support and Resilience. AIDS and Behavior, Ahead of Print

Vagni, M., Maiorano, T., Giostra, V., & Pajardi, D. (2020). Coping with COVID-19: Emergency Stress, Secondary Trauma and Self-Efficacy in Healthcare and Emergency Workers in Italy. Frontiers in Psychology, Ahead of Print

Weiner, L., Berna, F., Nourry, N., Severac, F., Vidailhet, P., & Mengin, A. C. (2020). Efficacy of an online cognitive behavioral therapy program developed for healthcare workers during the COVID-19 pandemic: the REduction of STress (REST) study protocol for a randomized controlled trial. Trials, 21(1), 1–10.

Xiao, H., Zhang, Y., Kong, D., Li, S., & Yang, N. (2020). The effects of social support on sleep quality of medical staff treating patients with coronavirus disease 2019 (COVID-19) in January and February 2020 in China. Medical science monitor: international medical journal of experimental and clinical research, 26, e923549–1.

Ye, Z., Yang, X., Zeng, C., Wang, Y., Shen, Z., Li, X., & Lin, D. (2020). Resilience, Social Support, and Coping as Mediators between COVID-19-related Stressful Experiences and Acute Stress Disorder among College Students in China. Applied Psychology: Health and Well-Being. Ahead of Print

Yörük, S., & Güler, D. (2020). The relationship between psychological resilience, burnout, stress, and sociodemographic factors with depression in nurses and midwives during the COVID-19 pandemic: A cross-sectional study in Turkey. Perspectives in Psychiatric Care. Ahead of Print

